# Self-reported word-finding complaints are associated with cerebrospinal fluid beta-amyloid and atrophy in cognitively normal older adults

**DOI:** 10.1101/2021.07.08.21260220

**Authors:** Maxime Montembeault, Stefan Stijelja, Simona M. Brambati, For the Alzheimer’s Disease Neuroimaging Initiative

## Abstract

**Background and Objectives:** Self-reported language complaints, and more specifically word-finding difficulties, are among the most frequent cognitive complaints in cognitively normal older adults (CN). The clinical significance of elevated self-reported word-finding complaints in CN is still a matter of debate. The present study aims at characterizing word-finding complaints in CN, establish their sociodemographic and psychological correlates, determine if they are predictive of lower levels of cerebrospinal fluid Aβ levels and finally, investigate if they are associated with brain atrophy in regions associated with naming impairments.

**Methods:** In this observational case-control study, 239 CN from the Alzheimer’s Disease Neuroimaging Initiative (ADNI) database were selected. All participants completed the self-reported version of the Everyday Cognition (ECog) questionnaire, as well as a lumbar puncture for Aβ and a MRI.

**Results:** Word-finding complaints were rated equally severe as a few other memory items and significantly more severe compared to all the other cognitive complaints. Ecog-Lang1 (Forgetting the names of objects) was not related to any demographic (age, sex, years of education) or psychological variable (depression-related symptoms, anxiety-related symptoms), while Ecog-Lang3 (Finding the right words to use in a conversation) was significantly negatively associated with years of education and positively associated with depression-related symptoms. Ecog-Lang1 severity significantly predicted CSF Aβ levels in CN, and this result remained significant even when controlling for all demographic and psychological variables as well as general level of cognitive complaint. Individuals with high Ecog-Lang1 complaints showed atrophy in the left fusiform gyrus and the left rolandic operculum in comparison to CN with no or low Ecog-Lang1 complaints.

**Discussion:** Overall, our results support the fact that word-finding complaints are significant in CN and should be taken seriously. They have the potential to identify CN at risk of AD and support the need to include other cognitive domains in the investigation of subjective cognitive decline.

## 1. Introduction

The amnestic form of Alzheimer’s disease (AD), in which episodic memory disturbances are predominant, is the most common form of the disease ^1^. Consequently, self-reported memory complaints among older adults have received considerable attention in recent years as a possible early marker of the disease ^2, 3^. This interest spawns, in part, from the fact that AD has a long prodromal phase where patients are either asymptomatic or have subtle cognitive complaints ^4^. Interestingly, subjective memory complaints in cognitively normal older individuals have been associated with the presence of amyloid ^2^ and brain atrophy ^3^.

Apart from memory deficits, a decline related to language performance, and most specifically word-finding difficulties, is also considered to be a very early sign of AD ^5-7^. Several studies have shown that language and more specifically word-finding complaints are among the most frequent complaints in cognitively normal older adults ^8-10^. In daily life, these word-finding complaints can manifest in different ways, such as forgetting the names of objects or people and having difficulties finding words in conversations. Even if the prevalence of word-finding complaints in healthy aging is elevated, the clinical significance of self-reported word-finding difficulties is still undetermined. General language complaints, which combine expressive (word-finding but other functions such as the ability to communicate thoughts efficiently for example) and receptive (comprehending language) abilities, have been investigated with conflicting results. On one hand, some studies suggest that language complaints might not be the most relevant predictors of dementia: they are not associated with AD pathology (Aβ and tau PET) ^11^ or enhanced risk of converting to MCI ^12^. On the other hand, a conflicting literature shows that higher language complaints are associated with CSF Aβ ^13-15^. In the only study investigating word-finding complaints specifically, Condret-Santi and colleagues found that subjective language complaints did not predict conversion to dementia in cognitively normal older adults ^8^.

No studies have investigated whether subjective word-finding complaints, specifically, can be considered an early indication of AD-related neuropathological and brain changes. In previous studies, subjective word-finding complaints assessment has been somewhat limited by considering language as a unitary domain or by the use of a single yes-no question. Also, sociodemographic and/or psychological factors might also have an impact on self-reported cognitive complaints. For instance, cognitive complaints were found to be correlated with sex and advanced age; women and older individuals displaying higher prevalence of subjective cognitive complaints ^16^. Language complaints, more specifically, have been related to depressive and anxiety-related symptoms ^10, 17^. Consequently, determining if word-finding difficulties are truly related to predictors of dementia might require accounting for these factors.

The present study aims at: 1) characterizing the frequency and severity of word-finding complaints in cognitively normal older adults (CN) compared to other language and cognitive domains; 2) identifying sociodemographic and psychological characteristics associated with word-finding complaints in CN; 3) determining if word-finding complaints can predict CSF Aβ levels and; 4) comparing gray matter volume in CN with varying levels of word-finding complaints. Based on the literature, we posit that 1) word-finding difficulties will be among the most frequent and severe cognitive complaints among CN; 2) age, sex, education, depression and anxiety might correlate with more severe word-finding complaints; 3) subjective language complaints will predict pathological levels of CSF a Aβ (even controlling for sociodemographic and psychological variables) and; 4) CN with significant language complaints will present gray matter atrophy in brain regions that are associated with naming and that are usually damaged in AD patients. Establishing the relationship between self-reported word-finding complaints and markers of Alzheimer’s disease might have important clinical implications. First, it is important to understand if cognitively normal older adults with a certain sociodemographic and/or psychological profile are more likely to experience them. Second, results would provide accurate information to older adults as to what they should be worried about. If word-finding complaints are shown to be related to AD biomarkers, this could contribute to early screening of older adults at higher risk of developing AD. Finally, it could highlight the relevance of incorporating non-memory complaints in the definition of subjective cognitive decline.

## 2. Methods

Data used in the preparation of this article were obtained from the Alzheimer’s Disease Neuroimaging Initiative (ADNI) database (adni.loni.usc.edu). The ADNI was launched in 2003 as a public-private partnership, led by Principal Investigator Michael W. Weiner, MD. The primary goal of ADNI has been to test whether serial magnetic resonance imaging (MRI), positron emission tomography (PET), other biological markers, and clinical and neuropsychological assessment can be combined to measure the progression of mild cognitive impairment (MCI) and early Alzheimer’s disease (AD). For up-to-date information, see www.adni-info.org.

### 2.1 Standard protocol approvals, registrations, and patient consents

The ADNI study was approved by an ethics standards committee on human experimentation at each institution in the United States and Canada. Written informed consent was obtained from all participants.

### 2.2 Data availability

The data used in this study are available by request to all qualified investigators on the ADNI database (adni.loni.usc.edu).

### 2.3 Participants

In the general ADNI inclusion criteria, subjects are deemed cognitively normal (CN) if: 1) they exhibited Mini-Mental State Exam (MMSE) scores between 24 and 30 (inclusive); 2) a Clinical Dementia Rating (CDR) score of 0; 3) scored above education adjusted scores on the delayed Paragraph Recall task from the Wechsler Memory Scale Logical Memory II (≥9 for 16 or more years of education, ≥5 for 8-15 years of education, ≥3 for 0-7 years of education); 5) were determined to be cognitively normal, based on an absence of significant impairment in cognitive functions or activities of daily living. Further information concerning enrollment and precise exclusion criteria can be accessed online (www.adni-info.org).

Study-specific inclusion criteria include 1) having available self-rated Everyday Cognition (ECog) questionnaire; and 2) having available cerebrospinal fluid (CSF) AD biomarkers. Therefore, 239 CN were included in the present study (Table 1). They were either enrolled in ADNI-GO (2009-2011) or ADNI-2 (2011-2017). The mean age was 73.11 ± 6.06 years (age range 56-89 years) and 47.3% (n=113) were men and 52.7% (n=126) were women. Mean years of education was 16.68 ± 2.46 years (education range 8-20 years).

**Table 1.** Demographic and psychological characteristics of the sample

| | Cognitively normal older adults(CN)<br>Mean ± SD | Range |
| --- | --- | --- |
| n | 239 | – |
| Male/Female (%) | 47.3/52.7 | – |
| Age | 73.1 ± 6.1 | 56.0 – 89.0 |
| Years of education | 16.7 ± 2.5 | 8.0 – 20.0 |
| CSF Amyloid beta (pg/ml) | 197.5 ± 50.1 | 82.7 – 303.0 |
| Ecog total score | 54.5 ± 13.0 | 39.0 – 102.0 |
| Geriatric Depression Scale (GDS) | 0.8 ± 1.1 | .0 – 6.0 |
| Anxiety item (Neuropsychiatric Inventory Examination) | 0.1 ± 0.7 | .0 – 8.0 |
| Mini-Mental State Exam (MMSE) | 29.1 ± 1.2 | 24.0 – 30.0 |

### 2.4 Clinical Assessment

The severity and frequency of word-finding difficulties were assessed using the self-reported Everyday Cognition (ECog) questionnaire. The ECog i.e. Everyday Cognition; ^18^ is a validated 39 items-scale probing an individual’s subjective complaints through 6 cognitive dimensions: memory (8 items), language (9 items), visuospatial (7 items), planning (5 items), organization (6 items) and divided attention (4 items). Responses range from 1 to 4 (1= no change or actually performs better than 10 years ago; 2 = occasionally performs the task worse than 10 years ago but not all of the time; 3 = consistently performs the task a little worse than 10 years ago; 4 = performs the task much worse than 10 years ago). Within the language dimension, 2 items are related to word-finding difficulties, namely Ecog-Lang1 (*“Forgetting the names of objects*”) and Ecog-Lang3 (“*Finding the right words to use in a conversation”)*. These two items represent our main variables of interest.

Individuals enrolled in ADNI were also required to perform a plethora of clinical and tests. The present study particularly considers two of them: Geriatric Depression Scale GDR; ^19^ to assess depression-related symptoms, and Neuropsychiatric Inventory Examination ^20^ to assess anxiety-related symptoms (anxiety item).

### 2.5 Biomarkers Collection

Amyloid-beta concentration (Aβ) was considered as the main biological proxy for probable AD dementia. The complete descriptions of the collection, transportation and analyses protocols are provided in the ADNI procedural manual at www.adni-info.org.

### 2.6 MRI acquisition and pre-processing

All subjects underwent the standardized MRI protocol of ADNI as described at http://www.loni.ucla.edu/ADNI/Research/Cores/index.shtml. Briefly, the ADNI protocol includes T1-weighted acquisition based on a sagittal volumetric magnetization-prepared rapid gradient echo sequence collected froma variety of MR systems with protocols optimized for each type of scanner. Representative imaging parameters were as follows: repetition time = 2300 ms; inversion time = 1000 ms; echo time = 3.5 ms; flip angle = 8°; field of view= 240 x 240 mm; and 160 sagittal 1.2-mm-thick slices and a 192 x 192 matrix yielding a voxel resolution of 1.25 x 1.25 x 1.2 mm, or 180 sagittal 1.2-mm-thick slices with a 256 x 256 matrix yielding a voxel resolution of 0.94 x 0.94 x 1.2 mm. The full details of the ADNI MRI protocol have been previously described (Jack, Bernstein, et al. 2008).

Image preprocessing was performed using CAT12 (http://www.neuro.uni-jena.de/cat/), which is an extension of SPM12 (Statistical Parametric Mapping 12, http://www.fil.ion.ucl.ac.uk/spm/software/spm12/), running on MATLAB (Mathworks, Natick, MA, USA). All T1-weighted images were corrected for bias (field inhomogeneities and noise). They were then segmented into gray matter (GM), white matter and cerebrospinal fluid (CSF) ^21^, spatially normalized and modulated. After pre-processing, all scans passed a visual check for artefacts and the automated CAT12 quality check protocol. The modulated and normalized gray matter images were then smoothed with a Gaussian kernel of 8mm FWHM.

### 2.7 Statistical analyses

All statistical analyses were performed using IBM SPSS Statistics 26 and R version 3.6. First, in order to compare the severity of Ecog-Lang1 and Ecog-Lang3 to the severity of all other cognitive complaints, a one-way repeated measures ANOVA with custom contrasts was carried. A Bonferroni-corrected statistical threshold of p ≤ .00066 was used for the seventy-six comparisons. Second, in order to identify demographic and psychological correlates of word-finding complaints in CN, Spearman rank correlations were used to establish the degree of association between ECog’s word-finding complaint items (ordinal variables) and selected demographic (age, sex, education) and psychological (depression-related symptoms, anxiety-related symptoms) variables. Third, we aimed to determine if word-finding complaints can predict CSF Aβ levels. One-way ANOVA tests were carried to determine if statistically significant differences were observable in CSF Aβ concentration between CN in each word-finding complaints’ levels. Multiple regression models were also fitted to analyze the ability of word-finding complaints to predict CSF Aβ levels when accounting for other covariates (demographic and psychological factors). Fourth, we aimed to compare gray matter volume in CN with varying levels of word-finding complaints. The VBM analysis was performed on smoothed GM images. Two one-way ANOVAs were performed to compare groups of CN according to their level of word-finding complaints (one for Ecog-Lang1 and one for Ecog-Lang3). Contrasts were set to compare GM volume between three groups: CN with no word-finding complaint, CN with occasional word-finding complaint and CN reporting consistent or much worse word-finding difficulties. Age, sex, handedness, total intracranial volume and scanner were included as nuisance covariates. Whole-brain statistical analyses were conducted at a statistical threshold of p < .05 familywise error (FWE), corrected for multiple comparisons was used.

## 3. Results

### 3.1 Severity and frequency of word-finding complaints in CN

Figure 1 presents the most severe cognitive complaints among our sample of 239 CN. When comparing the severity level of Ecog-Lang1 (*“Forgetting the names of objects*”) with all the other cognitive complaints, we found that it was equivalent to four other memory complaints (“Remembering a few shopping items without a list”, “Recalling conversations a few days later”, “Repeating stories and or questions”, “Remembering I have already told someone something”), but significantly higher than all other cognitive complaints (p ≤ .00066 Bonferroni-corrected). Ecog-Lang3 (“*Finding the right words to use in a conversation”)* was also significantly higher than most of the other cognitive complaints (p ≤ .00066 Bonferroni-corrected) and equivalent to three other memory complaints (“Remembering a few shopping items without a list”, “Remembering where I have placed objects”, “Remembering I have already told someone something”). Conversely, no other cognitive complaint was significantly higher than the two word-finding complaints.

**Figure 1.**
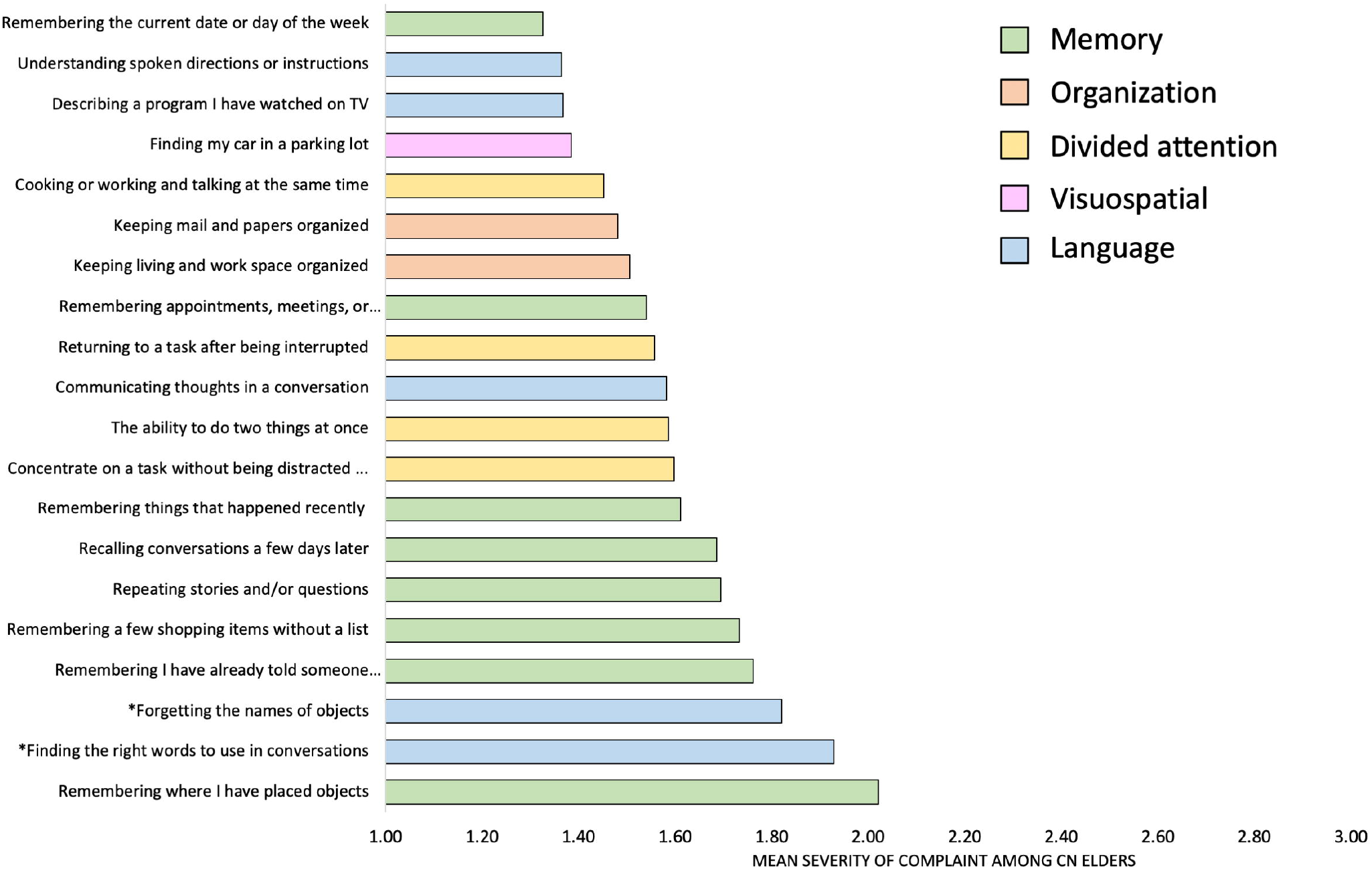
Levels of severity of cognitive complaints among cognitively normal older adults (20 top items out of the 39 ECog items).

In terms of frequency of word-finding complaints, while most CN report no change (36.0%) or being occasionally worse (47.2%) at forgetting the names of objects (Ecog-Lang1), 15.5% report being consistently a little worse and 1.3% report being consistently much worse. In terms of finding the right words to use in conversation (Ecog-Lang3), 31.0% report no change, 50.6% report being occasionally worse, 13.0% report being consistently a little worse and 5.4% report being consistently much worse. The distribution of responses per level of complaint for all the Ecog items is presented in Supplementary Table I. The most frequent subjective complaints across the 39 items of the Ecog, when considering the percent of CN who scored either 3 or 4, were the word-finding complaints (16%, 18%) as well as on two memory-related items (“Remembering a few shopping items without a list”:18% “Remembering where I have placed objects”:26%). Due to low response rates for the 4^th^ complaint level (4 = performs the task much worse than 10 years ago), the 3^rd^ and 4^th^ response options were merged for all the other analyses.

### 3.2 Demographic and psychological correlates of word-finding complaints in CN

Results for the Spearman rank correlations are presented in Table 2. Ecog-Lang1 was not significantly related with any demographic (age, sex, years of education) or psychological variable (depression-related symptoms, anxiety-related symptoms). Ecog-Lang3 was significantly negatively associated with years of education (*rho* = -.154; p = .017) and positively associated with depression-related symptoms (*rho* = .274; p = .000). There was a significant but moderate positive correlation between Ecog-Lang1 and Ecog-Lang3 (*rho* = 0.468; p = .000), which suggests that although they are related to some degree, they also capture different facets of word-finding complaints. Ecog-Lang1 and Ecog-Lang3 were therefore analysed separately for the remaining analyses.

**Table 2.** Spearman’s rank correlations between word-finding complaints and demographic/psychological variables.

|  | Sex | Age | Years of schooling | Geriatric Depression Scale (GDS) | Anxiety item (NPI) |
| --- | --- | --- | --- | --- | --- |
| Forgetting the names of objects (Ecog-Lang1) | -0.008 | 0.106 | -0.037 | 0.12 | 0.066 |
| Finding the right words to use in conversations (Ecog-Lang3) | 0.063 | -0.031 | -.154* | .274** | 0.068 |

\*\* . Correlation is significant at the 0.01 level (2-tailed).
\* . Correlation is significant at the 0.05 level (2-tailed).

### 3.3 CSF Aβ levels according to word-finding complaints severity levels

A statistically significant difference in CSF Aβ was found depending on the complaint severity of Ecog-Lang1 (*F*(2,236) = 5.044, p = .007; Figure 2A). The mean CSF Aβ level was 206.1 ± 50.9 for CN with no Ecog-Lang1 word-finding complaint, 198.6 ± 48.6 with occasional Ecog-Lang1 complaint and 176.3 ± 47.7 for CN who reported consistent or much worst Ecog-Lang1 complaint. A Tukey HSD post hoc test determined that the CN who reported having consistently or much more difficulty remembering the names of objects (severity level 3&4 combined) had significantly lower levels of CSF Aβ compared to the first (p = .005) and second level of complaint (p = .039).

**Figure 2.**
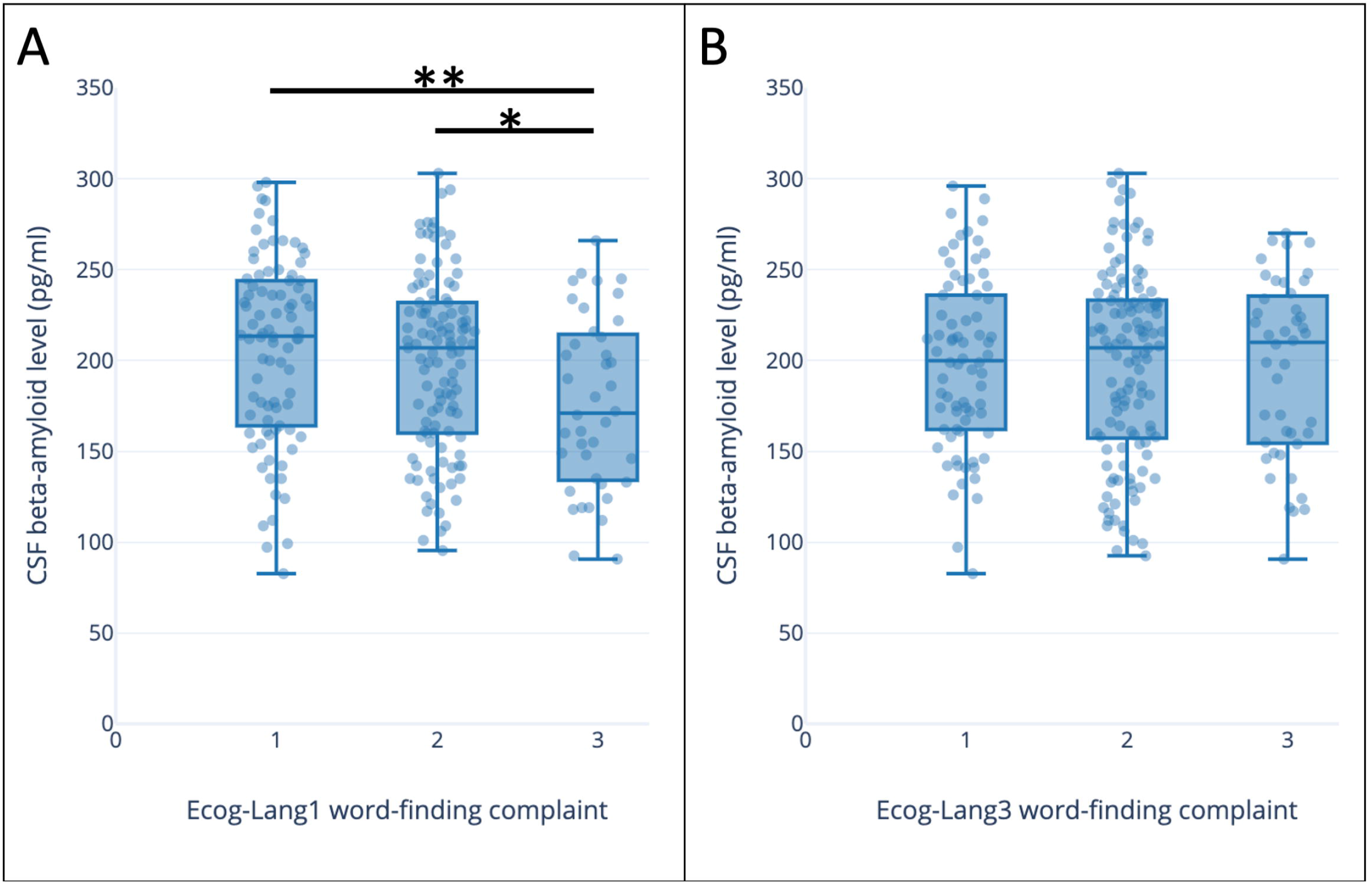
Aβ levels (pg/ml) according to severity of A) Ecog-Lang1 complaints (Forgetting the names of objects) B) Ecog-Lang3 complaints (Finding the right words to use in conversations) The values represent: 1 = No change or actually performs better than 10 years ago; 2 = Occasionally performs the task worse than 10 years ago but not all of the time; 3 = Consistently performs the task a little worse than 10 years ago or performs the task much worse than 10 years ago

When considering the second Ecog-Lang3, CSF Aβ levels did not differ significantly depending on the severity levels of the complaint (p = .903; Figure 2B).

### 3.4 Relation between word-finding complaints and CSF Aβ levels controlling for demographic and psychological factors

The results of our univariate analyses showed that the subjective complaint Ecog-Lang1 is related to CSF Aβ levels in CN. To increase confidence in these findings, we extended our previous analyses by including relevant control variables. Table 3 presents results of multiple regression models where CSF Aβ concentration is the dependant variable. The first model only included the predictor of interest (Ecog-Lang1), entered as a categorical variable. The second model was comprised of the total score on the Ecog (without Ecog-Lang1 and Ecog-Lang3), to assess if word-finding complaints are specifically associated with AD biomarkers, controlling for the overall degree of subjective cognitive complaints. Finally, a third model was fitted to control for demographic and psychological variables (sex, age, years of education, depression-related symptoms and anxiety-related symptoms). When all these covariates were included in the model, the most severe level of complaint predicted a statistically significant decrease of 26.906 pg/ml in CSF Aβ concentrations. Interestingly, total score on the Ecog (without items Ecog-Lang1 and Ecog-Lang3) did not significantly predict CSF Aβ levels (B = -0.025; p = .936).

**Table 3.** Regression analysis of predictors of CSF Aβ levels (Ecog-Lang1 as the main IV)

|  | **Model 1** | **Model 2** | **Model 3** |
| --- | --- | --- | --- |
| Ecog- Lang1(1/3) | *Ref* | *Ref* | *Ref* |
| Ecog- Lang1(2/3) | -7.501<br>(7.052) | -8.048<br>(7.301) | -5.106<br>(7.184) |
| Ecog- Lang1(3/3) | -29.816**<br>(9.431) | -31.555**<br>(11.106) | -26.906*<br>(10.902) |
| Ecog Total Score (Without<br>Lang1//Lang3) |  | .093<br>(.312) | -.013<br>(.318) |
| Sex |  |  | -15.885*<br>(6.606) |
| Age |  |  | -1.652**<br>(.526) |
| Years of Schooling |  |  | 2.022<br>(1.311) |
| Geriatric Depression Scale (GDS) |  |  | 2.191<br>(2.927) |
| Anxiety score |  |  | -4.037<br>(4.509) |
| Constant | 206.071***<br>(5.314) | 201.899***<br>(14.978) | 315.152***<br>(50.761) |

Standard errors in parentheses
\* p<0.05; \*\*p<0.01; \*\*\*p<0.001

### 3.5 Comparison of GM atrophy between individuals with varying levels of word-finding complaints

When considering Ecog-Lang1, CN reporting consistent or much worse word-finding difficulties had significant GM atrophy in the left fusiform gyrus (x = -29, y = -50, z = -12) and the left rolandic operculum (x = -44, y = -8, z = 11) in comparison to CN without word-finding complaint (p < .05 FWE; Figure 3). The reverse contrast did not show any significant difference. CN with occasional word-finding complaint did not significantly differ from any of the other groups either.

**Figure 3.**
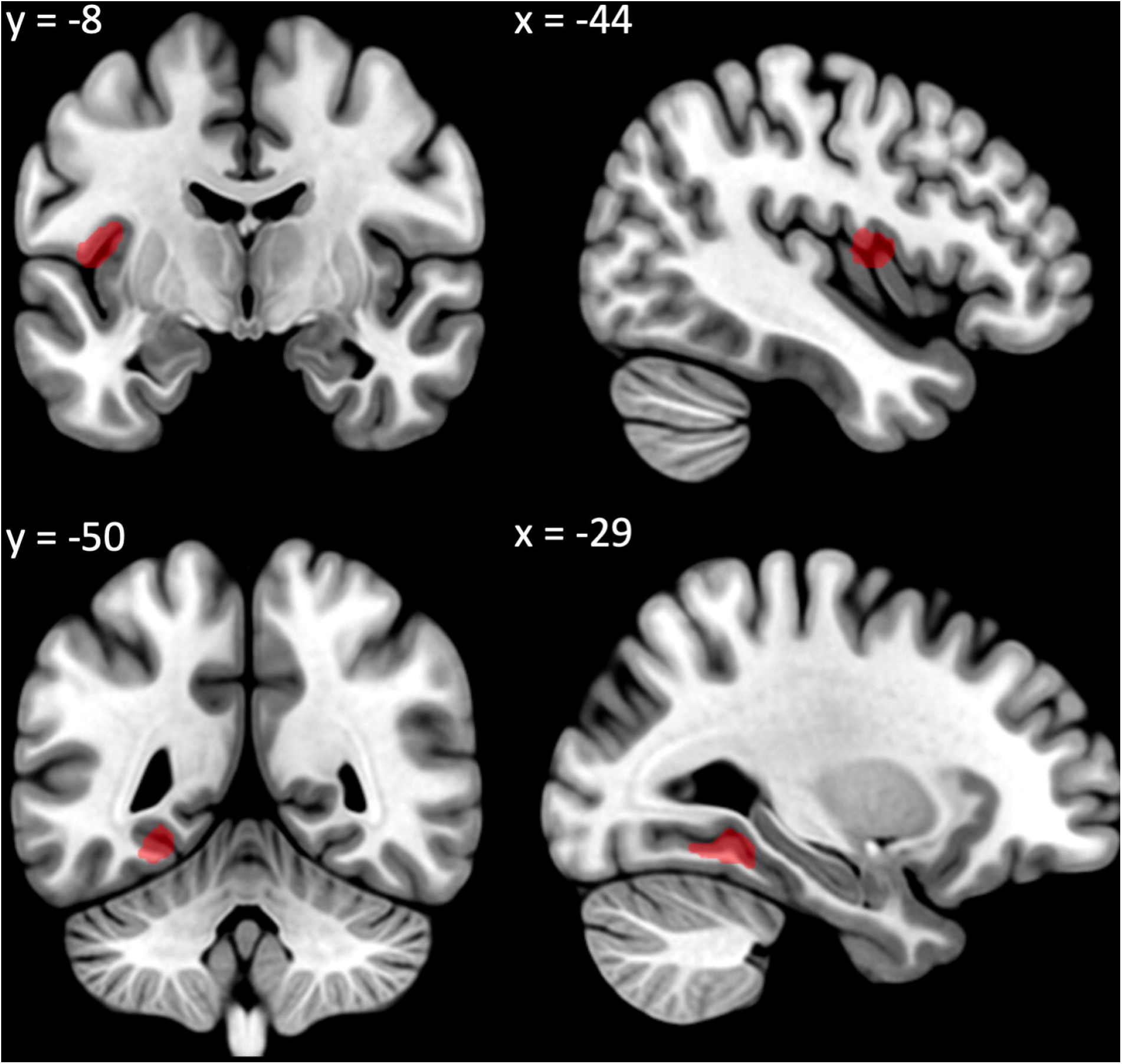
Decreased GM volume in participants with significant Ecog-Lang1 word-finding complaint (Forgetting the names of objects) vs. participants with no Ecog-Lang1 word-finding complaint (significant at p < .05 FWE corrected controlling for age, sex, handedness, scanner, total intracranial volume, presented at p < .001 uncorrected for display).

When considering Ecog-Lang3, GM volume did not differ significantly between groups with different levels of complaint.

## 4. Discussion

In this study, we demonstrated that word-finding complaints are amongst the most frequent and severe in CN. In this population, forgetting the name of objects was independent of main sociodemographic or psychological characteristics. Furthermore, it significantly predicted lower levels of CSF Aβ, even when controlling for sociodemographic variables, psychological measures and total cognitive complaints. Finally, CN who report consistently forgetting the names of objects show gray matter atrophy in the left fusiform gyrus and left rolandic operculum, in comparison to CN who don’t report this phenomenon or who only report it occasionally. Our results highlight the importance of word-finding complaints for the early screening of older adults at higher risk of developing AD.

### 4.1 Word finding complaints are amongst the most frequent and severe in CN

Our results show that word-findings complaints were as severe and frequent as other episodic memory complaints, and significantly more severe and frequent than all the other cognitive complaints (from the organization, divided attention and visuospatial domains). Approximately 50% of CN report occasional word-finding difficulties and 15-18% report consistent word-finding difficulties. These results suggest that CN are highly sensitive to word-finding changes, possibly due to the frequent use of language in daily life and the potential impacts of word-finding problems on social life. They are consistent with previous studies showing the elevated prevalence of such complaints ^8-10^. The current study adds to this literature by directly comparing word-finding complaints with complaints from other cognitive domains, therefore providing a complete profile of cognitive complaints in CN. Finally, word-finding complaints were significantly more severe than other types of language complaints, which highlight the importance of investigating word-finding complaints in isolation in CN. Otherwise, language complaints in CN would most likely be underestimated.

### 4.2 Forgetting the name of objects is independent of sociodemographic or psychological characteristics in CN

We were also interested in understanding if there are sociodemographic and/or psychological profiles that are associated with elders who are more likely to report experiencing word-finding difficulties. Our results suggest that age and sex are not related to word-findings complaints, contrary to previous studies suggesting that women and older individuals displayed higher prevalence of global subjective cognitive complaints ^16^. This has significant clinical implications: equivalent medical attention should be given to CN complaining of word-finding difficulties, no matter their age or sex. For the remaining characteristics, we obtained different associations depending on the word-finding complaint investigated (Ecog-Lang1 or Ecog-Lang3). Ecog-Lang1 (Forgetting the name of objects) was not related to education, depression-or anxiety-related symptoms. Its high prevalence in CN in our sample can therefore not be attributed to external factors such as psychological symptoms. Ecog-Lang3 (Finding the right words to use in conversations) was modestly correlated with education and depression-related symptoms, but not with anxiety-related symptoms. This suggests that this complaint is less specific and might be multifactorial. Previous studies have showed an association of subjective language complaints with psychological symptoms ^10, 17^. These partially conflicting results might be due to methodological considerations: significant depression-related symptoms are an exclusion criterion for the ADNI cohort and anxiety-related symptoms are not measured extensively (only with a general questionnaire investigating multiple neuropsychiatric symptoms, the NPI). Thus, although in our sample the association has not been observed, future studies should investigate word finding complaints in a more representative sample in which depression was not an inclusion criterion.

### 4.3 Word-finding complaints can help identify individuals with signs of AD pathology

Beyond underlining the high prevalence of word-finding complaints in healthy aging, one of the objectives of this study was to establish their clinical significance. Our study suggests that these complaints could serve to identify individuals presenting signs of pathology and who are at higher risk to develop AD dementia, before they show any objective signs of cognitive decline. In CN, greater self-reported difficulties to find the name of objects significantly predicted lower levels of CSF Aβ. Individuals with greater word-finding complaints also presented lower gray matter volumes in the left fusiform gyrus and the left rolandic operculum. Together, these results suggest that word finding complaints reported by CN can be the expression of early signs of AD pathology. This was specifically true for Ecog-Lang1 (Forgetting the names of objects) and not Ecog-Lang3 (Finding the right words to use in conversations). This can inform clinicians and researchers on the best way to interrogate patients about their word-finding difficulties: forgetting the names of objects appears very specific and is associated with AD biomarkers and brain atrophy, versus finding the right words to use in conversations which might be more multifactorial and isn’t predictive of such indicators.

Although word-finding complaints are specifically elevated in CN, beyond general language complaints, no study had previously investigated their associations with AD biomarkers and brain atrophy. Our results confirm that these complaints are worrying in older adults. Interestingly, CN who reported consistent or much worst Ecog-Lang1 complaint presented, on average, a level of CSF Aβ that is below the established cutoff of ≤192 pg/mL (176.3 ± 47.7; versus the other groups of patients with no or occasional Ecog-Lang1 complaints who were above this threshold) ^22^.. Our results are consistent with previous studies reporting an association between general language complaints and CSF Aβ in CN ^13-15^, further showing the relevance of word-finding complaints specifically. Even though Condret-Santi and colleagues found no increased risk of dementia in CN with word-finding complaints, they only assessed word-finding complaints using yes or no question. By investigating word-finding complaints using a continuous scale, we show that occasional word-finding difficulties is not associated with AD biomarkers or atrophy, while consistent word-finding difficulties is. Future longitudinal studies should investigate the relationship between continuous word-finding complaints and conversion to mild cognitive impairment or dementia in CN.

Our study is also the first to show that individuals with greater word-finding complaints present more brain atrophy in comparison to those with no or lower word-finding complains. This result was observed in the left fusiform gyrus and the left rolandic operculum, both regions implicated in language functions ^23-26^. Interestingly, the left fusiform gyrus is atrophied ^27^ and is implicated in naming ^23-25^ and semantic ^28^ impairments in AD patients. Interestingly, Pravatà and colleagues found that a year before AD diagnosis, MCI patients with naming difficulties showed distinct areas of greater GM loss in the left fusiform gyrus than MCI patients without naming difficulties. Our study suggest that these brain changes could happen even earlier, at a stage when individuals don’t present any cognitive impairment yet.

### 4.4 Word-finding complaints should be taken seriously and considered with other cognitive complaints in the screening of prodromal AD

Cognitive complaints are at the core of the diagnosis of subjective cognitive decline (SCD) and mild cognitive impairment ^4, 29^. Although these diagnostic criteria consider complaints from any cognitive domains, a majority of scientific studies on these populations have focused on episodic memory complaints ^4^. Because the available evidence for an association of preclinical AD with questions about memory functioning was the strongest when diagnostic criteria were written, a positive response to the question on subjective memory (and not other cognitive domains) has been proposed as an item of the SCD *plus* category (SCD with increased likelihood of the presence of preclinical AD) ^4^. Furthermore, it has been reported that most clinical tools to assess SCD focus on memory ^30^. More precisely, when considering all SCD assessment tools, 59% of items are related to memory while only 8% are related to language ^30^. Although our study was not specifically designed to investigate SCD, it suggests that word-finding complaints are central in older adults, that they are not the mere consequence of age, sex, education or psychological symptoms, and that it is rather associated to neuropathological and atrophy profiles that are characteristic of individuals at higher risk to develop AD. For this reason, word-finding complaints should be considered in the definition and screening of individuals with SCD.

### 4.5 Conclusion

Contrarily to previous studies suggesting that word-finding complaints are not worrying in older adults, our study actually suggests that these complaints should be taken seriously. This study highlights the importance of investigating word-finding subjective complaints in CN for the screening of AD, on top of other consistently investigated complaints such as memory-related complaints. This investigation should be at least specific to forgetting the names of objects and conducted using a continuous scale. Our results also advocate for the inclusion of other cognitive domains in the investigation of SCD, more precisely of word-finding complaints.

## Supporting information

eTable I

## Acknowledgements

MM is supported by postdoctoral funding from Fonds de Recherche en Santé du Québec (FRQS) and Canadian Health Institutes of Research (CIHR).

SMB is supported by research funding from Fonds de Recherche en Santé du Québec (FRQS) and Alheimer Society of Canada.

Data collection and sharing for this project was funded by the Alzheimer’s Disease Neuroimaging Initiative (ADNI) (National Institutes of Health Grant U01 AG024904) and DOD ADNI (Department of Defense award number W81XWH-12-2-0012). ADNI is funded by the National Institute on Aging, the National Institute of Biomedical Imaging and Bioengineering, and through generous contributions from the following: AbbVie, Alzheimer’s Association; Alzheimer’s Drug Discovery Foundation; Araclon Biotech; BioClinica, Inc.; Biogen; Bristol-Myers Squibb Company; CereSpir, Inc.; Cogstate; Eisai Inc.; Elan Pharmaceuticals, Inc.; Eli Lilly and Company; EuroImmun; F. Hoffmann-La Roche Ltd and its affiliated company Genentech, Inc.; Fujirebio; GE Healthcare; IXICO Ltd.; Janssen Alzheimer Immunotherapy Research & Development, LLC.; Johnson & Johnson Pharmaceutical Research & Development LLC.; Lumosity; Lundbeck; Merck & Co., Inc.; Meso Scale Diagnostics, LLC.; NeuroRx Research; Neurotrack Technologies; Novartis Pharmaceuticals Corporation; Pfizer Inc.; Piramal Imaging; Servier; Takeda Pharmaceutical Company; and Transition Therapeutics. The Canadian Institutes of Health Research is providing funds to support ADNI clinical sites in Canada. Private sector contributions are facilitated by the Foundation for the National Institutes of Health (www.fnih.org). The grantee organization is the Northern California Institute for Research and Education, and the study is coordinated by the Alzheimer’s Therapeutic Research Institute at the University of Southern California. ADNI data are disseminated by the Laboratory for Neuro Imaging at the University of Southern California.

