## Supplementary material for "Self-reported word-finding complaints are associated with cerebrospinal fluid beta-amyloid and atrophy in cognitively normal older adults": eTable I: eTable I.pdf

**eTable I** Frequency of subjective complaints among CN

| Type of Complaint | | Frequency | | | | | | | |
|---|---|---|---|---|---|---|---|---|---|
| | | 1 - Better or no change | | 2 - Occasionnally worse | | 3 - Constistently a little worse | | 4- Consistently much worse | |
| MEMORY1 | 1. Remembering a few shopping items without a list. | 116 | **49%** | 83 | **35%** | 28 | **12%** | 12 | **5%** |
| MEMORY2 | 2. Remembering things that happened recently (such as recent outings, events in the news). | 130 | **54%** | 77 | **32%** | 27 | **11%** | 5 | **2%** |
| MEMORY3 | 3. Recalling conversations a few days later. | 112 | **47%** | 93 | **39%** | 31 | **13%** | 3 | **1%** |
| MEMORY4 | 4. Remembering where I have placed objects. | 66 | **28%** | 111 | **46%** | 53 | **22%** | 9 | **4%** |
| MEMORY5 | 5. Repeating stories and/or questions. | 108 | **45%** | 102 | **43%** | 23 | **10%** | 6 | **3%** |
| MEMORY6 | 6. Remembering the current date or day of the week. | 178 | **74%** | 48 | **20%** | 9 | **4%** | 4 | **2%** |
| MEMORY7 | 7. Remembering I have already told someone something. | 94 | **39%** | 112 | **47%** | 29 | **12%** | 4 | **2%** |
| MEMORY8 | 8. Remembering appointments, meetings, or engagements. | 137 | **57%** | 79 | **33%** | 19 | **8%** | 4 | **2%** |
| **LANG1** | **1. Forgetting the names of objects.** | 86 | **36%** | 113 | **47%** | 37 | **15%** | 3 | **1%** |
| LANG2 | 2. Verbally giving instructions to others. | 179 | **75%** | 47 | **20%** | 12 | **5%** | 1 | **0%** |
| **LANG3** | **3. Finding the right words to use in conversations.** | 74 | **31%** | 121 | **51%** | 31 | **13%** | 13 | **5%** |
| LANG4 | 4. Communicating thoughts in a conversation. | 133 | **56%** | 78 | **33%** | 23 | **10%** | 5 | **2%** |
| LANG5 | 5. Following a story in a book or on TV. | 184 | **77%** | 46 | **19%** | 7 | **3%** | 2 | **1%** |
| LANG6 | 6. Understanding the point of what other people are trying to say. | 192 | **80%** | 42 | **18%** | 5 | **2%** | 0 | **0%** |
| LANG7 | 7. Remembering the meaning of common words. | 200 | **84%** | 35 | **15%** | 4 | **2%** | 0 | **0%** |
| LANG8 | 8. Describing a program I have watched on TV. | 165 | **69%** | 60 | **25%** | 14 | **6%** | 0 | **0%** |
| LANG9 | 9. Understanding spoken directions or instructions. | 163 | **68%** | 67 | **28%** | 7 | **3%** | 2 | **1%** |
| VISSPAT1 | 1. Following a map to find a new location. | 204 | **85%** | 27 | **11%** | 8 | **3%** | 0 | **0%** |
| VISSPAT2 | 2. Reading a map and helping with directions when someone else is driving. | 200 | **84%** | 32 | **13%** | 7 | **3%** | 0 | **0%** |
| VISSPAT3 | 3. Finding my car in a parking lot. | 158 | **66%** | 72 | **30%** | 7 | **3%** | 2 | **1%** |
| VISSPAT4 | 4. Finding my way back to a meeting spot in the mall or other location. | 181 | **76%** | 49 | **21%** | 7 | **3%** | 2 | **1%** |
| VISSPAT6 | 5. Finding my way around a familiar neighborhood. | 222 | **93%** | 16 | **7%** | 1 | **0%** | 0 | **0%** |
| VISSPAT7 | 6. Finding my way around a familiar store. | 220 | **92%** | 18 | **8%** | 1 | **0%** | 0 | **0%** |
| VISSPAT8 | 7. Finding my way around a house visited many times. | 231 | **97%** | 8 | **3%** | 0 | **0%** | 0 | **0%** |
| PLAN1 | 1. Planning a sequence of stops on a shopping trip. | 200 | **84%** | 35 | **15%** | 4 | **2%** | 0 | **0%** |
| PLAN2 | 2. The ability to anticipate weather changes and plan accordingly (i.e., bring a coat or umbrella) | 224 | **94%** | 14 | **6%** | 1 | **0%** | 0 | **0%** |
| PLAN3 | 3. Developing a schedule in advance of anticipated events. | 207 | **87%** | 30 | **13%** | 2 | **1%** | 0 | **0%** |
| PLAN4 | 4. Thinking things through before acting. | 179 | **75%** | 57 | **24%** | 3 | **1%** | 0 | **0%** |
| PLAN5 | 5. Thinking ahead. | 201 | **84%** | 36 | **15%** | 2 | **1%** | 0 | **0%** |

| | | | | | | | | | |
|---|---|---|---|---|---|---|---|---|---|
| ORGAN1 | 1. Keeping living and work space organized. | 145 | **61%** | 69 | **29%** | 23 | **10%** | 2 | **1%** |
| ORGAN2 | 2. Balancing the checkbook without error. | 199 | **83%** | 37 | **15%** | 3 | **1%** | 0 | **0%** |
| ORGAN3 | 3. Keeping financial records organized. | 190 | **79%** | 43 | **18%** | 5 | **2%** | 1 | **0%** |
| ORGAN4 | 4. Prioritizing tasks by importance. | 184 | **77%** | 47 | **20%** | 8 | **3%** | 0 | **0%** |
| ORGAN5 | 5. Keeping mail and papers organized. | 150 | **63%** | 65 | **27%** | 22 | **9%** | 2 | **1%** |
| ORGAN6 | 6. Using an organized strategy to manage a medication schedule involving multiple medications. | 213 | **89%** | 23 | **10%** | 3 | **1%** | 0 | **0%** |
| DIVATT1 | 1. The ability to do two things at once. | 125 | **52%** | 94 | **39%** | 14 | **6%** | 6 | **3%** |
| DIVATT2 | 2. Returning to a task after being interrupted. | 132 | **55%** | 85 | **36%** | 18 | **8%** | 4 | **2%** |
| DIVATT3 | 3. The ability to concentrate on a task without being distracted by external things in the environment. | 126 | **53%** | 87 | **36%** | 22 | **9%** | 4 | **2%** |
| DIVATT4 | 4. Cooking or working and talking at the same time. | 155 | **65%** | 63 | **26%** | 17 | **7%** | 4 | **2%** |
